# COVID-19 seroprevalence among healthcare workers of a large COVID Hospital in Rome reveals strengths and limits of two different serological tests: Seroprevalence to SARS-CoV-2 among healthcare workers: comparison by tests

**DOI:** 10.1101/2021.01.08.21249445

**Authors:** Giuseppe Vetrugno, Daniele Ignazio La Milia, Floriana D’Ambrosio, Marcello Di Pumpo, Roberta Pastorino, Stefania Boccia, Rosalba Ricci, Fabio De-Giorgio, Michela Cicconi, Federica Foti, Domenico Pascucci, Francesco Castrini, Elettra Carini, Andrea Cambieri, Maria Elena D’Alfonso, Gennaro Capalbo, Massimo Fantoni, Umberto Moscato, Domenico Staiti, Francesco Maria De Simone, Filippo Berloco, Maurizio Zega, Paola Cattani, Brunella Posteraro, Maurizio Sanguinetti, Patrizia Laurenti

**Author notes:** **Corresponding Author** (FDA).

## Abstract

In several hospitals worldwide, healthcare workers are currently at the forefront against coronavirus disease 2019 (COVID-19). Since Fondazione Policlinico Universitario A. Gemelli (FPG) IRCCS has been enlisted as a COVID hospital, healthcare workers deployed to COVID wards were separated from those with limited or no exposure, whereas administrative staff was destined to work-from-home. Between June 4 and July 3 2020, an investigation was carried out to evaluate seroprevalence of SARS-CoV-2 IgG antibodies among employees of the FPG using point-of-care (POC) and venous blood tests. Sensitivity, specificity and predictive values were determined with reverse-transcription polymerase chain reaction (RT-PCR) on nasal/oropharyngeal swabs as gold standard.

Four thousand, seven hundred seventy-seven participants were enrolled. Seroprevalence was 3.66% using the POC test and 1.19% using venous blood test, with a significant difference between the two (p < 0.05).

POC sensitivity and specificity were, respectively, 63.64% (95% confidence interval (CI): 62.20% to 65.04%) and 96.64% (95% CI: 96.05% to 97.13%), while those of the venous blood test were, respectively, 78.79% (95% CI: 77.58% to 79.94%) and 99.36% (95% CI: 99.07% to 99.55%). Among low-risk population, point-of-care’s predictive values were 58.33% (positive) and 98.23% (negative) whereas venous blood test’s were 92.86% (positive) and 98.53% (negative). In conclusion, point-of-care tests have low diagnostic accuracy, while venous blood tests seem to show an overall poor reliability.

## Introduction

On December 2019, a cluster of a unknown acute respiratory illness occurred in Wuhan city, Hubei Province in China, and rapidly spread to other areas in the following months [1,2]. The responsible agent was identified by the Chinese Centre for Disease Control and Prevention (CCDC) on January 7 2020 and was subsequently named severe acute respiratory syndrome coronavirus 2 (SARS-CoV-2). The disease was later named COVID-19 by the World Health Organization (WHO) [3]. Due to the widespread global transmission of COVID-19 and the high rate of contagiousness, the WHO declared COVID-19 to be a pandemic on March 11 2020 [3]. Early in the SARS-CoV-2 outbreak, several healthcare workers (HCWs) have been infected while providing care to patients with COVID-19 [4,5,6]. Identification and isolation of infected and potentially infectious HCWs is indeed relevant to protect them and their families and, besides, may prevent onward transmission to patients and colleagues as well as reduce the risk of healthcare-associated outbreaks [5]. In Italy, COVID-19 cases increased rapidly since February 23 2020 with SARS-CoV-2 spreading mostly in northern regions, particularly in the Lombardy region, where on June 4 2020 the number of COVID-19 cases were 89,526 (38.26% of total cases in Italy). Although in the Lazio region, at the same date, COVID-19 cases were 7764, the Fondazione Policlinico Universitario A. Gemelli (FPG) IRCCS—a large teaching University hospital in Rome enlisted as a COVID hospital—treated 553 COVID-19 patients, 133 of them in the intensive care unit (ICU). Based on these evidences and in accordance with the Lazio region, [7] the FPG launched a seroprevalence investigation to assess potential contagion sources among FPG HCWs. Previous studies already discussed data on HCWs seroprevalence over different countries worldwide, indicating that isolation protocols, hygiene standards and personal protective equipment (PPE) may prevent high levels of nosocomial transmission [8-17].

Current diagnostic tests for COVID-19 fall into two main categories: molecular tests detecting SARS-CoV-2 RNA and serological tests detecting anti-SARS-CoV-2 immunoglobulins (Igs; i.e. IgG/IgM) [18]. The reverse-transcription polymerase chain reaction (RT-PCR) molecular test, usually performed on nasal/oropharyngeal swab (NOS) samples, is considered the reference standard for COVID-19 diagnosis [19]. However, this test has long turnaround times (it takes over 2 to 3 hours to generate results) and requires certified laboratories, expensive equipment and trained technicians to operate. Limitations include potential false negative results and precarious availability of test materials [18,19]. Conversely, serological tests have been proposed as an alternative to RT-PCR in cases of acute SARS-CoV-2 infection [20]. They are, in addition, cheaper and easier to implement in laboratory diagnostics for SARS-CoV-2, especially in a point-of-care (POC) format. A clear advantage of these tests over RT-PCR is that they can identify individuals previously infected by SARS-CoV-2, even if they never underwent testing while acutely ill [18]. Considering their short time of appearance from the SARS-CoV-2 infection onset, viral specific IgG and IgM antibodies could indicate an ongoing infection [19]. In this regard, population-based sero-epidemiological surveys, especially in healthcare settings, quantifying the proportion of individuals with anti-SARS-CoV-2 antibodies, may be very helpful [21]. The aim of this study was to assess seroprevalence of SARS-CoV-2 specific IgG antibodies among HCWs of the FPG, using serological tests, which rely on venous or capillary blood sampling. The sensitivity and specificity of both tests, namely venous blood and POC test, respectively, were evaluated in comparison with RT-PCR results on NOS samples from FPG HCWs.

## Materials and methods

### Study participants and design

This cross-sectional study consisted in a seroprevalence survey between June 4 2020 and July 3 2020, which enrolled via a hospital e-mail system, on a voluntary basis, medical, non-medical HCWs and administrative staff (AS) of the FPG (Table 1). The study was approved by the FPG Ethics Committee (number ID 3253) and participants signed an informed consent before their inclusion in the study. Both venous blood and POC SARS-CoV-2 serological tests were offered to each participant and performed in dedicated blood-drawing areas in compliance with COVID-19 safety regulations. In case of positivity to at least one test, participants underwent NOS sampling for RT-PCR SARS-CoV-2 RNA detection to assess the actual infection status [7]. Unlike venous blood testing, POC testing was performed by trained clinical staff composed by public health residents and student nurses.

**Table 1.** General characteristics of enrolled participants.

| Participants' characteristics | N (%) |
| --- | --- |
| Age class |  |
| <36 years | 1414 (30.38) |
| 36-45 years | 1127 (24.21) |
| 46-55 years | 1240 (26.64) |
| >56 years | 874 (18.76) |
| Sex |  |
| Males | 1655 (34.65) |
| Females | 3122 (65.35) |
| Professional category |  |
| Medical doctors | 829 (17.35) |
| Nurses | 1481 (31.00) |
| Medical residents | 552 (11.56) |
| Other HCWs | 1059 (22.17) |
| Administrative staff | 474 (9.92) |
| External workers | 207 (4.33) |
| Others/unclassified | 175 (3.66) |
| COVID care |  |
| Yes | 736 (15.41) |
| No | 4041 (84.96) |
| Total participants | 4777 (100) |

Medical and non-medical HCWs were categorized into two groups whether they had assisted or had not assisted COVID-19 patients, in the period between March 9 2020 (date of the first COVID-19 patients in our hospital) and June 4 2020 (date of seroprevalence survey initiation). For predictivity analysis, which requires to consider the prevalence of the studied population, we used AS as further comparison group, because this was a group with low seroprevalence. This is due to the fact that these participants were less exposed to COVID-19 infection than HCWs and many of them had been in work-from-home for two days a week from March 9 2020.

As above mentioned, participants, which tested positive for SARS-CoV-2 specific antibodies, with at least one of the serological tests used in the study (see below), were sampled for NOS testing [22] within 48 hours after positive serological test results were available. RT-PCR testing on NOS samples was performed using the Seegene Allplex(tm) 2019-nCoV assay, and a positive result (i.e., a Ct less than 40) for at least one of two viral targets (i.e., RdRP and N genes) indicated the presence of SARS-CoV-2 RNA.

### Detection of anti-SARS-CoV-2 antibodies in blood samples

Two serological tests were used to detect SARS-CoV-2 specific antibodies in participants’ blood samples. In venous blood tests, venous blood samples were subjected to an enzyme-linked immunosorbent assay (ELISA) marketed by Euroimmun (Lübeck, Germany; www.euroimmun.com) for SARS-CoV-2 IgG detection [23]. In POC tests, capillary blood samples were directly subjected to the AllTest™ 2019-nCoV IgG/IgM Rapid Test Cassette assay lateral-flow chromatographic immunoassay for SARS-CoV-2 IgG/IgM detection [19,24].

### Detection of COVID-19 infection among healthcare workers

The collection of respiratory tract specimen, through NOS, to confirm COVID-19 status, was routinely performed at least once on 5270 HCWs, who met the following criteria: symptomatic participants; contact without adequate PPE with a COVID-19 case; HCWs employed in a ward with COVID-19 patients (swab performed monthly); HCWs employed in a ward with COVID-19 patients (swab performed bi-monthly); and HCWs not employed in a COVID-19 ward (swab performed quarterly) [19].

### Sensitivity and specificity of serological tests

The sensitivity and specificity of the ELISA-based venous blood and POC tests were assessed using RT-PCR assay as the diagnostic gold standard. In particular, we included only RT-PCR results from NOS samples obtained from the participants at least 30 days before. This allowed to define the seroconversion time for each participant with a positive serological result. As comparison group for predictive value analysis, we considered all participants enrolled in the study with at least one negative RT-PCR molecular test and only AS with at least one negative RT-PCR molecular test, because HCWs are a group more exposed to COVID-19 infection risk than the general population. Besides, AS seems to have a lower risk of positivity to SARS-CoV-2, similarly to general population, than most exposed groups, such as HCWs or quarantined persons who had been exposed to SARS-CoV-2 [25].

### Statistical analysis

Descriptive analysis was performed for sex, age, professional category and wards of the HCWs. Difference between proportions was evaluated with the two proportions z test. Seroprevalence was calculated, separately, for test on venous sampling and POC test. Seroprevalence for test on venous sampling was estimated as the proportion of individuals who had a positive result in the IgG in the immunoassay. Furthermore, for tests on capillary blood, both seroprevalence for IgM and IgG was estimated, as the proportion of individuals who had positive result in the corresponding band of the POC test.

Besides, we also re-estimated the sensitivity and specificity of the POC test using the immunoassay as a reference. The accuracy of the capillary versus the venous test was evaluated with sensitivity, specificity and predictive values with 95% confidence intervals (CI) [18]. All the seroprevalence was stratified for professional category, age and wards in which they worked during the COVID-19 emergency.

The difference between positivity to one of the serological tests and positivity to the RT-PCR on NOS samples was estimated through Pearson’s chi-squared test. The Spearman Rank test (Bonferroni adjusted) was used to evaluate the correlation between the anti-SARS-CoV-2 IgG assay in capillary blood versus the same titre in venous blood. Furthermore, Cronbach’s alpha was evaluated. In general, significant reliability values for Cronbach’s alpha are to be considered those > 0.70.

Statistical analyses were carried out using a software for the construction of the general basic dataset (Microsoft Excel for Mac Version 16.35), and specific for the inferential statistical analysis (Stata Corp 4905 Lakeway, College Station, USA Stata / IC 14.2 for Mac (64-bit Intel), Revision 29 Jan 2018).

## Results

Of the 7889 eligible participants, 4888 (62%) responded to the invitation and 111 of them refused to participate. Therefore, 4777 participants were enrolled in the seroprevalence investigation. Of the enrolled participants, 295 participants expressed consent to the venous blood test only, 83 expressed consent to the POC test only, and 4399 expressed consent for both. Table 1 reports the general characteristics of the participants. Their mean age was 43.11 years (SD ± 11.51), age data was missing for 122 participants. Females accounted for 65.35% (3122/4777) of the total; 31% (1481/4777) of the participants were nurses and 17.35% (829/4777) medical doctors. Around 15% (736/4777) of the participants was employed in COVID-19 wards.

The rate of positivity for SARS-CoV-2 RNA detected on NOS samples was 0.85% (45/5270). Conversely, POC seroprevalence was 3.66% (164/4482) and venous blood test seroprevalence was 1.19% (56/4694). Stratified results are shown in Table 2. Participants tested at least once for SARS-CoV-2 RT-PCR detection in NOS samples were 3538. Of these, 3184 had a RT-PCR result at least 30 days before serological testing and these results were considered in the accuracy analysis of serological tests.

**Table 2.** Prevalence of SARS-CoV-2 infection based on the three tests, overall and stratified by age, sex, professional category, direct assistance to COVID-19 patients or not (COVID care Yes/No).

|  | Participants with a positive result by each test (%) |
| --- | --- |

| Participants' characteristics | Venous blood test (%) | POC test (%) | RT-PCR test (%) |
| --- | --- | --- | --- |
| Overall | 56 (1.19%) | 164 (3.66%) | 45 (0.85%) |
| Age class |  |  |  |
| <36 years | 16 (1.15%) | 4 (3.75%) | 16 (0.86%) |
| 36-45 years | 11 (1.62%) | 47 (3.57%) | 10 (0.91%) |
| 46-55 years | 8 (0.82%) | 28 (2.96%) * | 8 (0.62%) |
| >56 years | 8 (1.40%) | 43 (4.98%) * | 10 (0.89%) |
| Sex |  |  |  |
| Males | 23 (0.01%) | 45 (0.02%) | 18 (0.01%) |
| Females | 33 (0.01%) | 119 (0.04%) | 27 (0.01%) |
| Professional category |  |  |  |
| Medical doctors | 20 (2.43%) <sup>1</sup> | 30 (3.80%) <sup>3</sup> | 14 (1.34%) |
| Nurses | 16 (1.10%) <sup>2</sup> | 59 (4.25%) <sup>4</sup> | 15 (0.77%) |
| Medical residents | 4 (0.74%) <sup>2</sup> | 20 (3.74%) <sup>3</sup> | 6 (0.66%) |
| Other HCWs | 11 (1.06%) <sup>2</sup> | 39 (3.93%) <sup>1</sup> | 9 (0.75%) |
| Administrative staff | 2 (0.43%) <sup>2</sup> | 10 (2.21%) <sup>6</sup> | 0 (0.00%) |
| External workers | 0 (0.00%) <sup>2</sup> | 1 (0.52%) <sup>5</sup> | 0 (0.00%) |
| Others/unclassified | 3 (1.83%) <sup>2</sup> | 5 (3.82%) <sup>3</sup> | 1 (1.49%) |
| COVID care |  |  |  |
| Yes | 10 (1.38%) | 33 (4.98%) * | 12 (1.10%) |
| No | 46 (1.16%) | 131 (3.43%) | 33 (0.79%) |
| <sup>1</sup> vs nurses, medical residents, other HCWs, administrative staff, external workers<br><sup>2</sup> vs medical doctors only<br><sup>3</sup> vs external workers only<br><sup>4</sup> vs external workers and administrative staff<br><sup>5</sup> vs all other categories<br><sup>6</sup> vs medical doctors, nurses, external workers<br>* p<0.05 |  |  |  |

Considering all participants enrolled, the POC test showed a sensitivity of 63.64% (95% CI, 62.20% to 65.04%) and a specificity of 96.64% (95% CI, 96.05% to 97.13%), whereas the venous blood test showed a sensitivity of 78.79% (95% CI, 77.58% to 79.94%) and a specificity of 99.36% (95% CI, 99.07% to 99.55%). Conversely, considering only as a comparison group, the POC test showed a sensitivity of 63.64% (95% CI, 59.10% to 67.93%) and a specificity of 97.79% (95% CI, 95.90% to 98.85%), whereas the venous blood test showed a sensitivity of 78.79% (95% CI, 74.89% to 82.22%) and a specificity of 99.57% (95% CI, 98.36% to 99.92%). Positive (PPV) and negative predictive values (NPV) for the POC and venous blood test and stratification of PPV, NPV, sensitivity and specificity with both AS and total participants as comparison group are shown in Table 3.

**Table 3.** Diagnostic parameters for POC and venous blood tests stratified with both AS and total participants as comparison groups.

|  | Administrative comparison group |  |  | Total participants comparison group |  |  |
| --- | --- | --- | --- | --- | --- | --- |
|  |  | Lower limit | Upper limit |  | Lower limit | Upper limit |
| Venous blood test |  |  |  |  |  |  |
| Sensitivity | 78.79% | 74.90% | 82.23% | 78.79% | 77.59% | 70.95% |
| Specificity | 99.57% | 98.37% | 99.92% | 99.36% | 99.07% | 99.56% |
| Negative predictive value | 98.53% | 96.92% | 99.34% | 99.85% | 99.68% | 99.93% |
| Positive predictive value | 92.86% | 90.15% | 94.88% | 46.43% | 44.99% | 47.87% |
| POC test |  |  |  |  |  |  |
| Sensitivity | 63.64% | 59.11% | 67.94% | 63.64% | 62.21% | 65.04% |
| Specificity | 97.79% | 95.91% | 98.85% | 96.64% | 96.06% | 97.13% |
| Positive predictive value | 58.33% | 53.74% | 62.79% | 8.54% | 7.74% | 9.40% |
| Negative predictive value | 98.23% | 96.46% | 99.15% | 99.81% | 99.63% | 99.92% |

Pearson’s chi-squared test showed a significant difference between the POC and venous blood test results (p < 0.05).

Out of 4683 observations, Spearman’s rho was 0.1052 with a p-value < 0.00.

Cronbach’s alpha showed a low concordance (scale reliability coefficient: 0.4477 p < 0.05). Of the 7889 eligible participants, 45 HCWs were COVID-19 known cases before the beginning of seroprevalence survey. Of these cases, 33 were enrolled in the survey and 26 of 33 resulted positive for SARS-CoV-2 IgG with the venous blood test. None of the participants resulting positive with the venous blood or POC test, consequently tested for SARS-CoV-2 infection by RT-PCR, had detectable SARS-CoV-2 RNA in their NOS samples.

## Discussion

Our results show that the rates of SARS-CoV-2 infection and serological positivity in different work categories are consistent with the low spread of SARS-CoV-2 in the FPG, which differs from the national surveillance data reporting a seroprevalence of 2.5% [26]. Furthermore, the seroprevalence for HCWs ranged between 1.6% and 14.6% in several studies [8-17].

Besides, we found a slight difference (but not statistically significant) in the seroprevalence determined by venous blood test between HCWs, who assistance or not assisted in COVID-19 wards: this is in accordance with another study reporting a higher seroprevalence rate than the one reported in our study [27]. The highest seropositivity rates (with venous blood test) among different worker categories were observed for medical doctors. Although this finding is not of immediate interpretation, a possible reason could be their exposure to high risk procedures (i.e., oral intubation, reanimation, clinical examination). Overall, we observed low differences (but not statistically significant) in seropositivity rates between sex and age categories.

Our study shows that the specificity was high for venous blood but not for POC test. Conversely, using RT-PCR assay as the diagnostic gold standard, venous blood test sensitivity might meet the criteria for screening tests, unlike POC test sensitivity. NPV were acceptable for both tests, whereas the PPV was acceptable only for the venous blood test, as shown in Table 3. A systematic review and meta-analysis by Bastos et al. [18] revealed that the current evidence does not support continued use of existing POC tests for COVID-19 serology. On one hand, our study shows a high PPV for venous blood serological testing among a low-risk population (i.e., AS) of hospital staff, on the other hand, this finding may not be the case for a medium/high-risk population (i.e., the entire working community of a COVID hospital).

We also observed that, among the subset of participants who tested positive with the venous blood or POC test and who were consequently tested for SARS-CoV-2 infection by RT-PCR, none of these participants had detectable SARS-CoV-2 RNA in their NOS samples, thus confirming that the serological test is not useful to diagnose COVID-19. Of 33 COVID-19 known cases tested for SARS-CoV-2 IgG in the venous blood, only 26 (78.8%) resulted positive, thus highlighting disagreement with recent observations [25] and confirming uncertainties about infections that occurred more than five weeks before the tests [20]. Our evidence suggests a decrease of the antibody titre, which in turn could corroborate the hypothesis of a non-persistent immunity and ultimately justify a possible re-infection with SARS-CoV-2 [28]. Furthermore, our sample, while small in size, is composed mainly of mild or asymptomatic cases (only 2 of 33 cases were admitted to ICU). As proposed by Burgess et al., [29] severity of illness is linked to the magnitude of serological responses. This association is also suggested by the experience gathered from other coronaviruses [30].

Our large teaching hospital gave us the opportunity to enroll a conspicuous sample size. Furthermore, even if some of the published studies involved larger numbers, our study is the first stratifying the sample by risk. From the beginning of the pandemic in Italy, the hospital directorate decided to distinguish HCWs who work in dedicated COVID-19 wards from those with limited or no exposure (i.e., working in non-COVID wards), and allowed work-from-home for AS. We assume that the last group has the same risk of the general population as AS was employed in work-from-home, during the Italian lockdown period.

Our study had several limitations. The sample was not drawn randomly and the estimation of the seroprevalence was also subject to other potential sampling bias, due to the voluntarily enrolling procedure. Moreover, the sensitivity of the serological test could be biased because it depends on the test time from the onset of disease. Samples collected from infected individuals outside the time window of antibody response could produce false negatives and, therefore, the observed seroprevalence in our study could potentially underestimate the true prevalence of the disease. Another limitation of the study could consist in the COVID-19 case detection strategy. In fact, to ascertain the exact number of COVID-19 cases among HCWs, a systematic and periodic (ideally every 14 days) screening with RT-PCR from NOS samples should be performed. However, this was not sustainable during the emergency due to lack of resources.

Owing to the cross-sectional design of this study, the dynamic changes of antibody titre in infected individuals over time were not evaluated.

Further studies, especially with long-term follow-up, will be needed in the future to assess the value of serological tests, considering their major public health implications.

## Data Availability

All relevant data are within the manuscript and its Supporting Information files.

## Acknowledgement

Lucia Zaino, Carmen Nuzzo, Nicola Nicolotti, Raffaele Pignataro, Davide Cammarata, Ilaria Amadio, Giovanna Guidi, Priscilla Emili, Grazia Morandotti, Carolina Castagna, Martina Sapienza for their kind support; Franziska Michaela Lohmeyer for language editing; Region Lazio Government for the supply of venous collection kits.

